# Cross-Modality Synthetic Data Augmentation using GANs: Enhancing Brain MRI and Chest X-ray Classification

**DOI:** 10.1101/2024.06.09.24308649

**Authors:** Kunaal Dhawan, Siddharth S. Nijhawan

## Abstract

Brain MRI scans and chest X-ray imaging are pivotal in diagnosing and managing neurological and respiratory diseases, respectively. Given their importance in diagnosis, the datasets to train the artificial intelligence (AI) models for automated diagnosis remain scarce. As an example, annotated chest X-ray datasets, especially those containing rare or abnormal cases like bacterial pneumonia, are scarce. Conventional dataset collection methods are labor-intensive and costly, exacerbating the data scarcity issue. To overcome these challenges, we propose a specialized Generative Adversarial Network (GAN) architecture for generating synthetic chest X-ray data representing healthy lungs and various pneumonia conditions, including viral and bacterial pneumonia. Additionally, we extended our experiments to brain MRI scans by simply swapping the training dataset and demonstrating the power of our GAN approach across different medical imaging contexts. Our method aims to streamline data collection and labeling processes while addressing privacy concerns associated with patient data. We demonstrate the effectiveness of synthetic data in facilitating the development and evaluation of machine learning algorithms, particularly leveraging an EfficientNet v2 model. Through comprehensive experimentation, we evaluate our approach on both real and synthetic datasets, showcasing the potential of synthetic data augmentation in improving disease classification accuracy across diverse pathological conditions. Indeed, the classifier performance when trained with fake + real data on brain MRI classification task shows highest accuracy at 85.9%. Our findings underscore the promising role of synthetic data in advancing automated diagnosis and treatment planning for pneumonia, other respiratory conditions, and brain pathologies.

## III. Introduction

Chest X-ray imaging stands as a cornerstone in the diagnosis and management of respiratory diseases, with pneumonia representing a prevalent yet challenging condition. The accurate interpretation of chest X-ray images is vital for timely diagnosis and treatment planning. However, the availability of annotated chest X-ray datasets, particularly encompassing rare or abnormal cases such as bacterial pneumonia, remains scarce [1]. This scarcity poses significant challenges to the development and evaluation of machine learning algorithms for automated diagnosis, hindering progress in this critical healthcare domain [5].

Moreover, conventional approaches to dataset collection and labeling are often labor-intensive, time-consuming, and expensive. The process involves expert radiologists manually annotating large volumes of images, leading to high labor costs and potential privacy concerns due to the need to access patient data. Additionally, obtaining labeled data for rare or abnormal cases like bacterial pneumonia can be particularly challenging, further exacerbating the data scarcity issue [2].

To address these challenges, the utilization of Generative Adversarial Networks (GANs) [3] has emerged as a promising solution. GANs enable the generation of synthetic medical images that closely resemble real patient data, thereby augmenting existing datasets and mitigating the limitations imposed by data scarcity [3]. By leveraging GANs, researchers can create diverse and realistic synthetic chest X-ray images representing both healthy lungs and various pathological conditions, including pneumonia.

In this paper, we propose a GAN architecture tailored specifically for the generation of synthetic chest X-ray data, representing both healthy lungs and various pneumonia conditions (viral and bacterial). Our approach aims to address the scarcity of labeled data and alleviate the burden of data collection and labeling, while also addressing privacy concerns associated with accessing patient data.

Furthermore, we demonstrate how synthetic data generation can facilitate the development and evaluation of machine learning algorithms for chest X-ray classification [4]. We highlight the role of artificial intelligence (AI) in automating the diagnosis of pneumonia by training an EfficientNet v2 [6] model on a mix of real and synthetic data, emphasizing the potential benefits of utilizing synthetic data to train and validate these deep learning models. Additionally, we conduct a thorough evaluation of model performance on both real and synthetic datasets, demonstrating the potential of synthetic data augmentation in improving disease classification accuracy. We evaluate our approach on both viral and bacterial pneumonia cases, where viral pneumonia is common and bacterial pneumonia is rare, showcasing the effectiveness of synthetic data in enhancing classification accuracy across diverse pathological conditions.

Moreover, we extend our experiments to include brain MRI scans by simply swapping the training dataset, achieving similar positive trends. This extension demonstrates the robustness of our proposed GAN approach in different medical imaging contexts, further highlighting the power of synthetic data generation in advancing machine learning applications across healthcare.

## IV. Brief literature review

GANs are increasingly being used to aid in creating and assembling training data to provide the annotated datasets to help in development of medical diagnosis models using convolutional neural networks (CNN). [1] describes how the medical datasets are often limited in size due to data annotation and privacy concerns, thus calling for the need to increase the dataset size without collecting or annotating more real data.

The recent applications of CNNs in radiology have led to advancements and use case across each of the four categories: classification, segmentation, detection, and others [6]. In medical image analysis, classification with deep learning utilizes target lesions depicted in medical images, and these lesions are classified into two or more classes, for example, benign or malignant. For segmentation, one of the common ways is to use a CNN classifier for calculating the probability of the image being an organ or anatomical structure. In this approach, the segmentation process is divided into two steps; the first step is the construction of the probability map of the organ or anatomical structure using CNN and image patches, and the second is a refinement step where the database of images and the probability map are utilized.

On the other hand, the basic Generative Adversarial Networks (GAN) model is made up of the generator and discriminator networks, in addition to the input vector [13]. This way, GAN is known to be able to “learn” the generative model of a given data distribution with excellent performance. This overcomes the issue of less labelled and unbalanced classification data being available, which doesn’t lend itself to medical diagnosis. In digital image processing, GAN can help with high-resolution image generation from low-resolution images.

Even using small datasets [14], different state-of-the-art GANs have shown the capability of generating usable image dataset for training CNNs, which while using these synthetic datasets outperform the baseline CNN model using only real data for training by up to 15.3% with respect to the F1 score, especially for datasets containing less than 100 images. The success of GANs also emanates from their sensitivity to tweaking the hyperparameters while the image synthesis is happening [2] to provide usable images for diagnosis.

[15] conducted a multi-GAN and multi-application study to assess the benefits of GANs in medical imaging. They did this by testing various GAN architectures (ranging from the basic DCGAN to more advanced style-based GANs) and established that the top-performing GANs can generate realistic-looking medical images by FID standards, though the capability to reproduce the full richness of medical datasets is still evolving. They further describe how style based GANs introduce multiple innovations to gradually train the GAN with different resolutions. These come with more developed generator, which includes adaptive instance normalization blocks (AdaIN) that enables the noise to be injected at every level of the network and use an 8-layer MLP mapping function. Literature also describes these GAN training innovations or “tricks”, such as label smoothing, feature matching, and differentiable augmentation of the images.

## V. Methodology

The proposed research automatically learns the underlying structure and distribution of real-world data, chest X-ray images in our case, and generates new, synthetic images with similar characteristics and abnormalities. This task is performed by employing a Generative Adversarial Network (GAN) which utilizes two separate neural network modules, termed as generator (G) and discriminator (D). G and D compete with each to generate synthetic instances of data which can be perceived as real ones [3]. The workflow is showcased in figure 1. From a given dataset containing real chest X-ray scans of various patients, we first form various pre-processing steps to ensure the images are in a suitable state for training the GAN. We apply normalization by thresholding and dilating each image to remove excess noise, and then crop each instance with dynamic coordinates to further denoise the image. Finally, we reshape each image to fit the dimensions 112 × 112 as a standard across the entire dataset.

**Figure 1.**
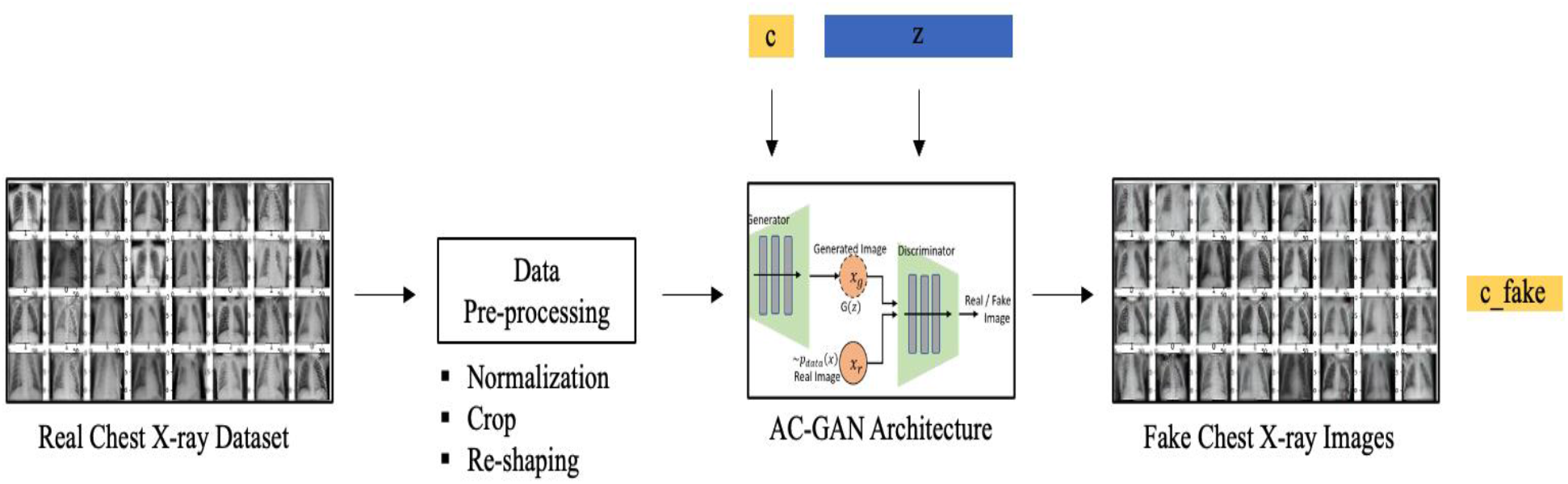
Workflow of the proposed Algorithm

After pre-processing, we apply a latent vector z containing random noise to each image sample across each class label c (healthy, viral pneumonia, bacterial pneumonia). Next, we train the proposed GAN model based on AC-GAN [9] to generate chest X-ray images across 3 classes defined above, along with their respective class labels c_fake. Usage of AC-GAN based architecture stabilizes the training process and facilitates generation of high-quality images while learning a representation which is independent of the class label.

For the modality of brain MRI scans, we use the exact same workflow and swap chest X-ray images with brain MRI images.

### V.1. Dataset

We use a dataset titled ‘Chest X-Ray Images (Pneumonia) [7], available on Kaggle as an open source database of 5,863 X-Ray images in JPEG format across two categories, normal lungs and pneumonia-ridden lungs. The diseased lungs contain two types of abnormalities, namely, bacterial pneumonia and viral pneumonia (samples shown in figure 2.). Existing statistics [8] reveal that viral pneumonia is more prevalent among patients, while bacterial pneumonia accounts for a smaller percentage of the total pool.

**Figure 2.**
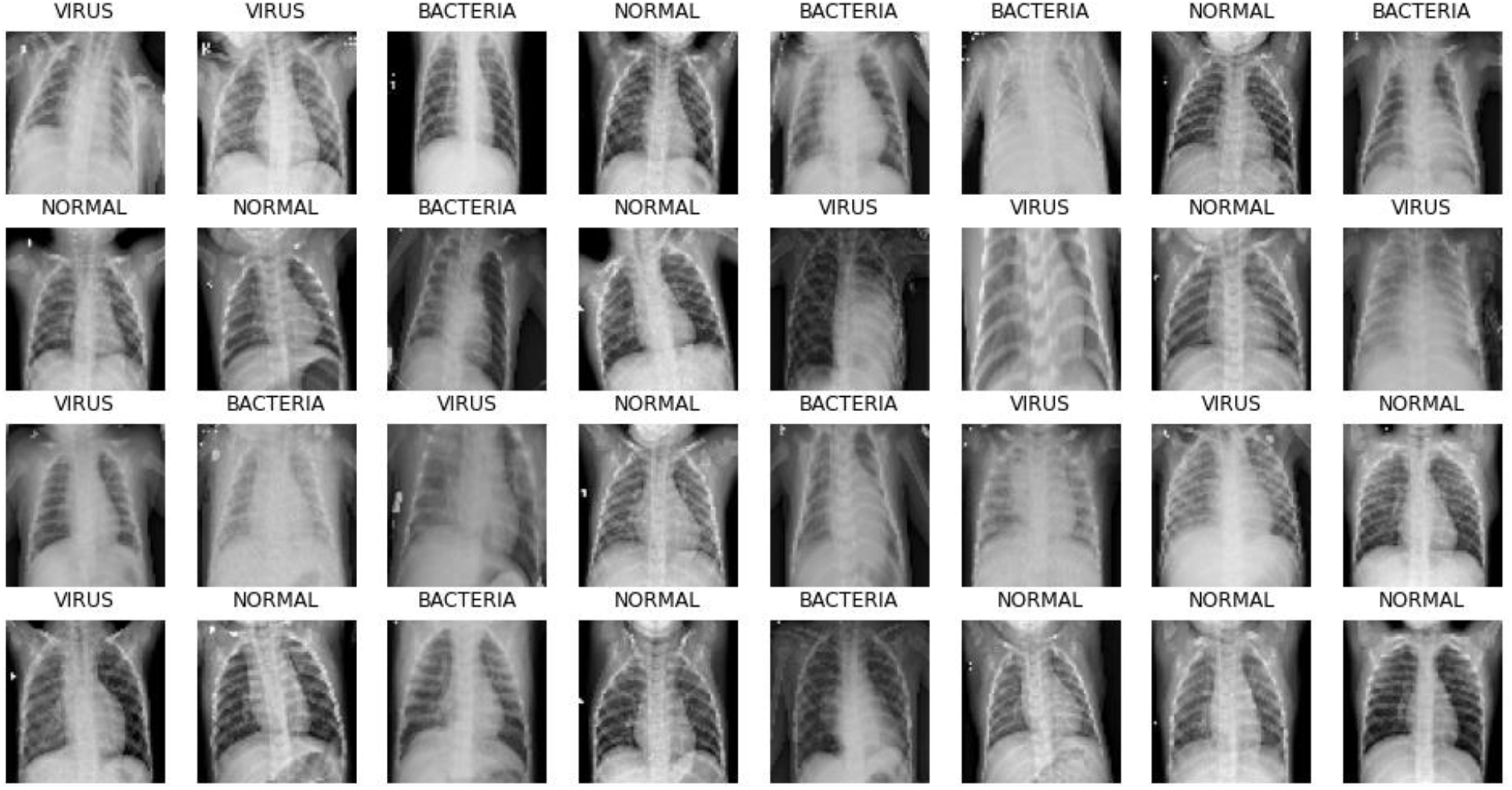
Real image samples of chest X-ray across 3 classes: normal, bacteria and virus

For the task of generating synthetic brain MRI images, we use another dataset titled ‘Brain Tumor Dataset’ [12], available on Kaggle. It is also an open source collection of medical imaging data, originally designed for the purpose of brain tumor detection and classification. The dataset contains 5,266 MRI scans of the human brain across two categories: positive (containing tumors) and negative (healthy brain without tumors). Some samples are illustrated in figure 3.

**Figure 3.**
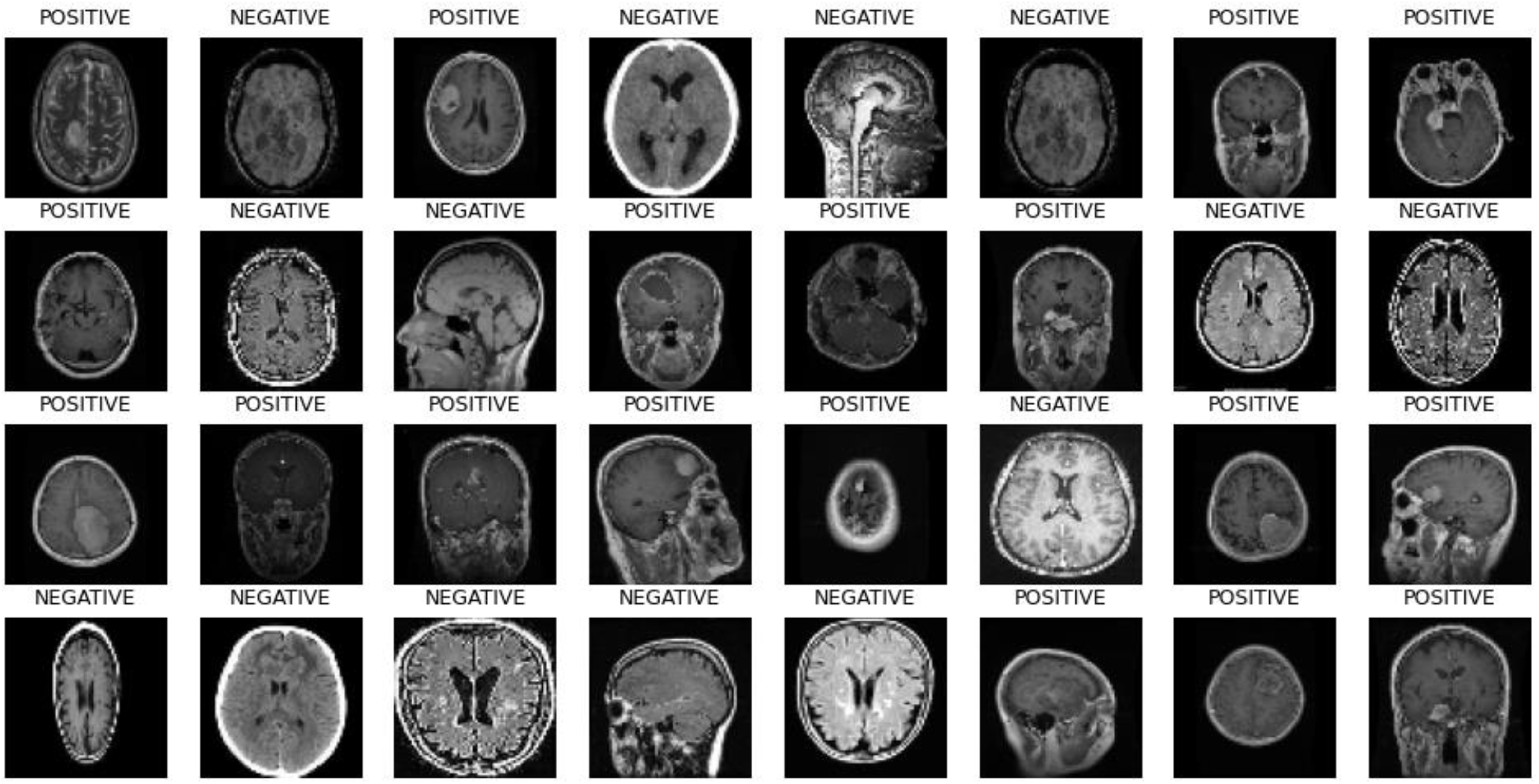
Real image samples of brain MRI across 2 classes: positive and negative

### V.2. Data Pre-processing

Chest X-ray scans can induce intensity averages falsely due to various distortions introduced to the system while scanning the subject. This condition may lead to the algorithm falsely identifying a healthy X-ray as diseased, therefore, pre-processing stage is extremely important. We perform normalization of the entire image which basically acts as a function to reduce the intensity values within a range suitable for the GAN to learn the spatial characteristics of the X-ray. This step sets the mean intensity close to 0 and standard deviation close to 1. Image is also denoised using several image processing techniques like erosions, averaging and dilations. Further, we remove the black portions of the image background by performing crop operation, whose coordinates are derived by capturing the largest contour of torso cross-section in the scans. Since our network takes an input of shape 112 × 112, we re-shape each image to this dimension before feeding it to our GAN for training and generation. We apply the same pre-processing pipeline to the brain MRI scans.

### V.3. AC-GAN Architecture

The Auxiliary Classifier Generative Adversarial Network (AC-GAN) [10] framework, proposed by [11], extends the traditional GAN architecture to integrate class information into the training process. This augmentation enables the generation of samples conditioned on specific class labels, thereby enhancing the controllability and diversity of the generated data distribution.

In the AC-GAN framework, the generator network is augmented to accept both random noise vectors sampled from a standard Gaussian distribution and class labels as input. By conditioning the generation process on class labels, the generator learns to produce class-specific features in the synthesized samples. This conditioning mechanism facilitates the generation of diverse samples corresponding to different classes. Similarly, the discriminator network in AC-GAN is modified to predict both the authenticity of the samples (real or synthetic) and their corresponding class labels. By jointly optimizing the discriminator for both tasks, distinguishing real from synthetic samples and correctly classifying the samples into their respective classes, the AC-GAN framework encourages the discriminator to learn discriminative features for each class. Its ability to generate class-conditioned samples makes it particularly suitable for tasks where controlling specific attributes of the generated samples is essential, such as generating synthetic medical images representing different pathological conditions.

In our proposed GAN architecture shown in figure 4, we draw inspiration from the AC-GAN framework to incorporate class conditioning into both the generator and discriminator networks. This enables us to generate synthetic chest X-ray images representing both healthy lungs and various pneumonia conditions (viral and bacterial), thereby enhancing the diversity and realism of the generated data.

**Figure 4.**
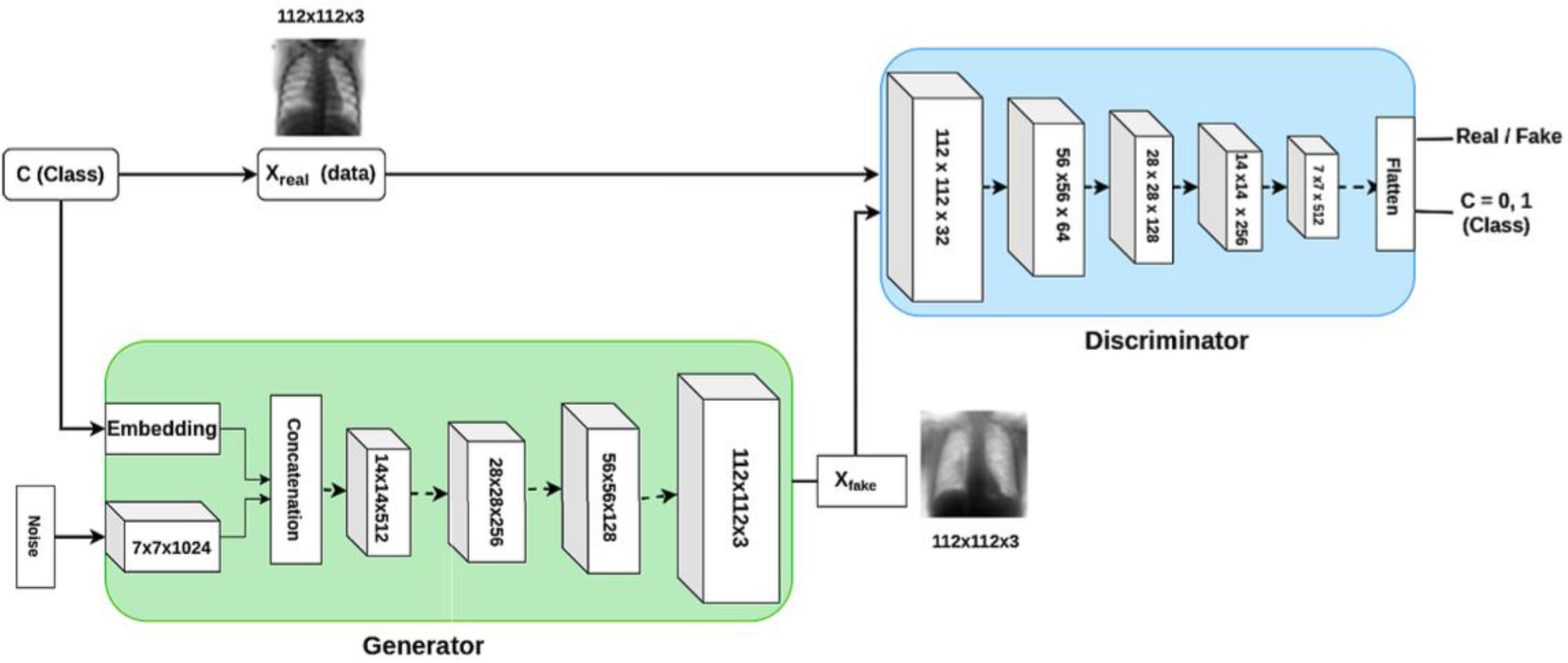
AC-GAN Architecture [9]

The generator G takes a latent vector of noise and class label as input, to output a single 112 × 112 × 3 image. An embedding of dimension 50 is generated using the class label, which is further passed through a 7 × 7 dense layer with linear activation. The noise vector is passed through a 1024 × 7 × 7 dense layer to generate multiple copies of low-resolution fake images. These are then concatenated and passed through 4 transpose convolutional layers, upsampling them at each stage. We employ batch normalization after each of the first 3 transpose convolution layers along with ReLU activation. The discriminator D consists of multiple convolutional layers followed by batch normalization, Leaky ReLU activation and a dropout layer. The input is downsized at each stage. The output contains 2 layers, a sigmoid to predict real / fake images, and another softmax to output class prediction probabilities.

### V.4 Implementation Details

Training an AC-GAN involves alternating between updating the parameters of the generator and discriminator networks through backpropagation. During each training iteration, the generator aims to produce realistic samples that can deceive the discriminator into classifying them as real, while the discriminator endeavors to accurately classify the samples into their corresponding classes and distinguish between real and synthetic samples. We split the dataset outlined above into 3 sets and use it for training, validation and testing. The model architecture is implemented in the TensorFlow library. The model is trained for a total of 32000 epochs with a batch size of 32 images. We use Root Mean Square Propagation (RMSProp) as the optimizer function with a learning rate of 1e-4. The end-to-end design and development was done in Jupyter Notebook hosted on Kaggle Cloud ML Engine.

## VI. Experimental results

In this section, we visualize the synthetic data generated using AC-GAN in the form of chest X-ray images across 3 classes: ‘Normal’, ‘Bacteria’, and ‘Virus’. Then, to quantitatively evaluate the effectiveness of using synthetic data to train an image classification model EfficientNet v2, we compute several metrics with the model trained on real data only, fake data only, and a mix of both real and fake data. To compare the efficacy of chest disease classification, we compute the classification accuracy. We also compute precision, which indicates the accuracy of the positive predictions made by the model. To evaluate the ability of the model to correctly identify positive instances, we also compute recall value. In the real world, the amount of images belonging to the normal class are more compared to diseased classes of bacteria and viruses. Therefore, this leads to an issue of class imbalance. We mitigate this issue by computing F1-score which provides a balance between precision and recall and is particularly useful when the class distribution is imbalanced.

Figure 5 reports the visual results of images generated using our proposed AC-GAN belonging to ‘Normal’ class, i.e., healthy lungs. Similarly, figure 6 and 7 showcase the images belonging to class ‘Bacteria’ and ‘Virus’ respectively. Visual inspection of the generated images reveals realistic representations of chest X-rays across different classes. For the class “Normal,” the generated images exhibit characteristic features of healthy lungs, including clear lung fields and well-defined structures. These images closely resemble authentic chest X-rays, demonstrating the AC-GAN’s ability to capture and reproduce normal anatomical structures faithfully. Similarly, for the classes “Bacteria” and “Virus,” the generated images display distinct pathological features associated with bacterial and viral pneumonia, respectively. These features include infiltrates, consolidations, and opacities indicative of the respective pneumonia types. The generated images exhibit variations in lesion size, shape, and distribution, reflecting the diversity of pathological manifestations observed in clinical practice.

**Figure 5.**
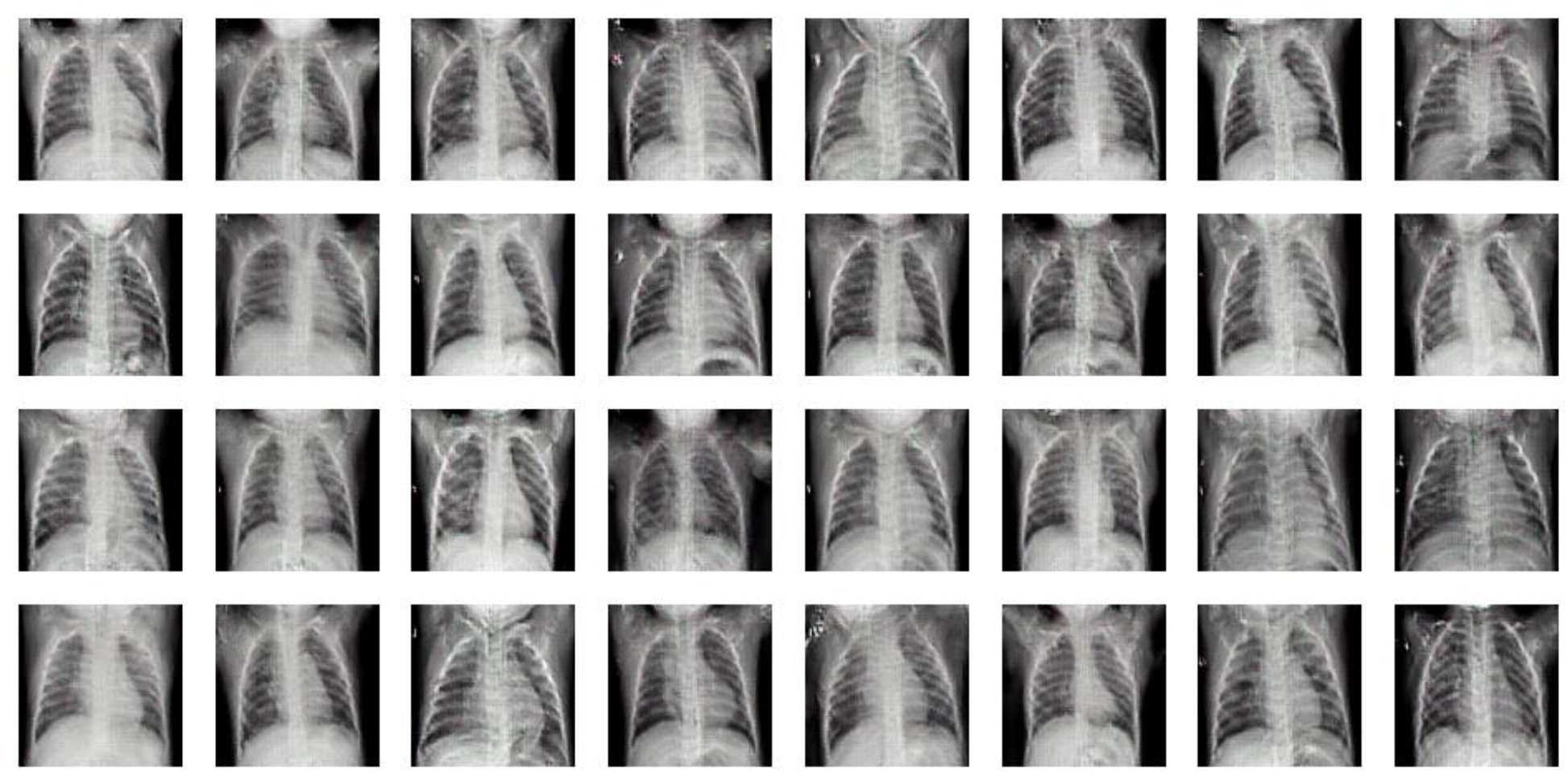
Images of class ‘Normal’ generated by proposed AC-GAN

**Figure 6.**
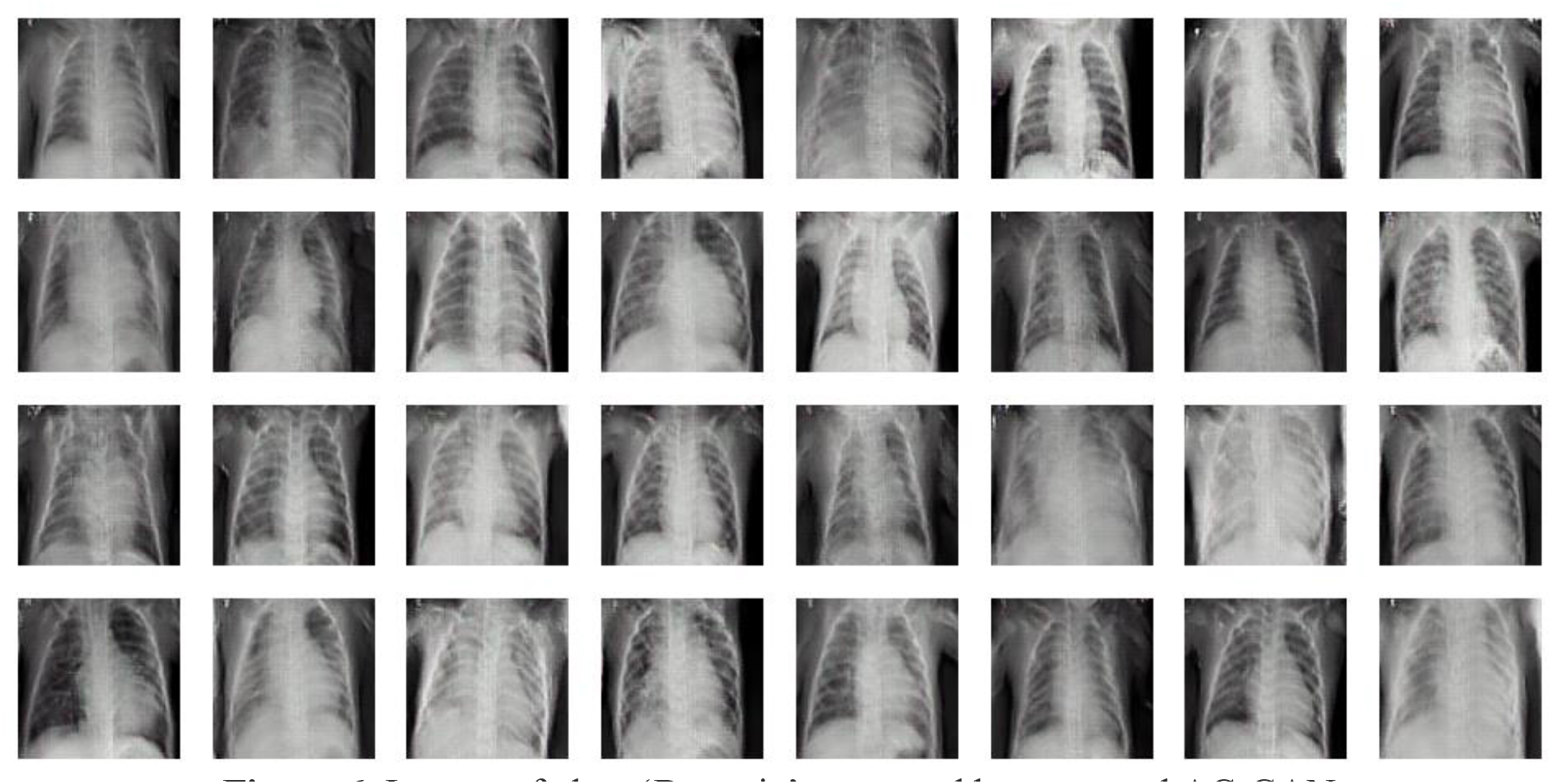
Images of class ‘Bacteria’ generated by proposed AC-GAN

**Figure 7.**
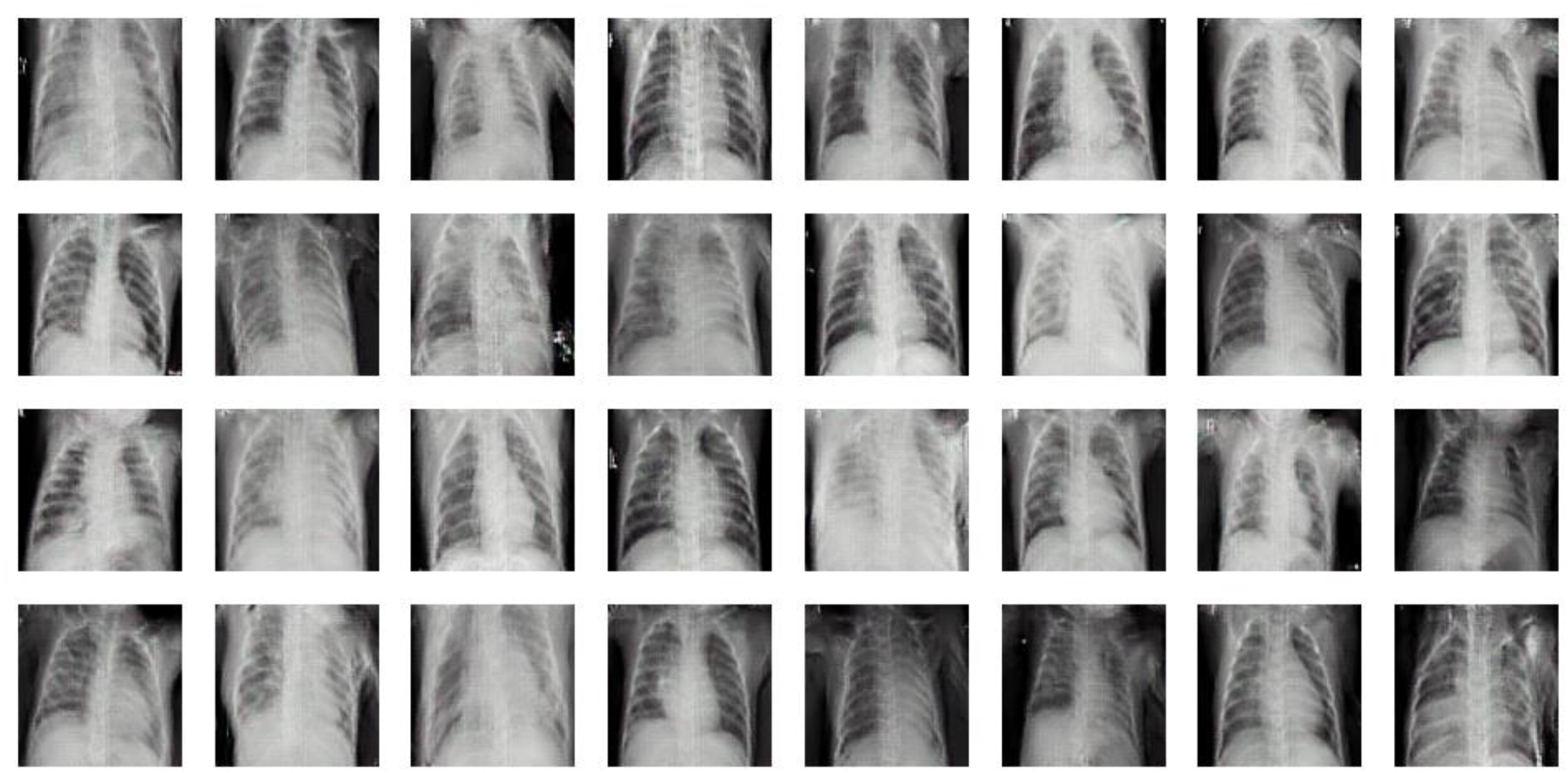
Images of class ‘Virus’ generated by proposed AC-GAN

Overall, the quality of the generated chest X-ray images is impressive, showcasing the AC-GAN’s capability to generate diverse and realistic images representing both normal and pathological conditions. This qualitative assessment is complemented by quantitative evaluation metrics reported in table I.

**Table I.** Classifier performance when trained with real, fake, and fake + real data on chest.

| Dataset | Accuracy %<br>↑ | Precision ↑ | Recall ↑ | F1-score ↑ |
| --- | --- | --- | --- | --- |
| **Real** | 70.66 % | 0.67 | 0.63 | 0.57 |
| **Fake** | 75.36 % | **0.89** | 0.75 | 0.77 |
| **Fake + real** | **79.83 %** | 0.84 | **0.77** | **0.93** |

The findings reveal that training solely with limited real data resulted in the lowest performance across all evaluation metrics as it exhibited the least accuracy, precision, recall, and F1-score compared to the other training scenarios. In contrast, incorporating synthetic data alongside real data yielded notable improvements in performance metrics. The combined dataset achieved the highest accuracy of 79.83%, along with a peak recall score of 0.77 and an impressive F1-score of 0.93, showcasing its ability to effectively identify positive instances across all classes. While the precision metric peaked at 0.89 for the classifier trained solely on fake data, the combined dataset demonstrated competitive precision scores while achieving superior accuracy. These results highlight the importance of leveraging synthetic data augmentation techniques to augment limited real datasets, thereby enhancing the robustness and effectiveness of image classifiers in medical image analysis tasks such as chest X-ray classification.

We further analyze the training progress of the image classifier through loss versus epoch curves for the three training scenarios, as showcased in figure 8. In the case of training with real data only, the loss versus epoch curve reveals a typical pattern of overfitting, where the training loss significantly decreases while the validation loss remains higher. This discrepancy indicates that the model is excessively fitting to the training data, resulting in poor generalization performance on unseen validation data. Conversely, training solely on fake data exhibits a smoother training process, with both training and validation losses converging gradually without signs of overfitting. This phenomenon is expected, as the synthetic data augment the training set, providing a more diverse and representative sample for the model to learn from compared to the limited real data. For the combined dataset comprising both real and fake data, the training converges rapidly, reflecting the effectiveness of leveraging synthetic data to complement real data. However, after a certain number of epochs, the loss versus epoch curve shows signs of overfitting, where the training loss continues to decrease while the validation loss starts to increase. To mitigate overfitting, early stopping is employed, terminating the training process before the model’s performance on the validation set deteriorates significantly. This ensures a fair evaluation of the model’s performance and prevents it from being overly biased towards the training data.

**Figure 8.**
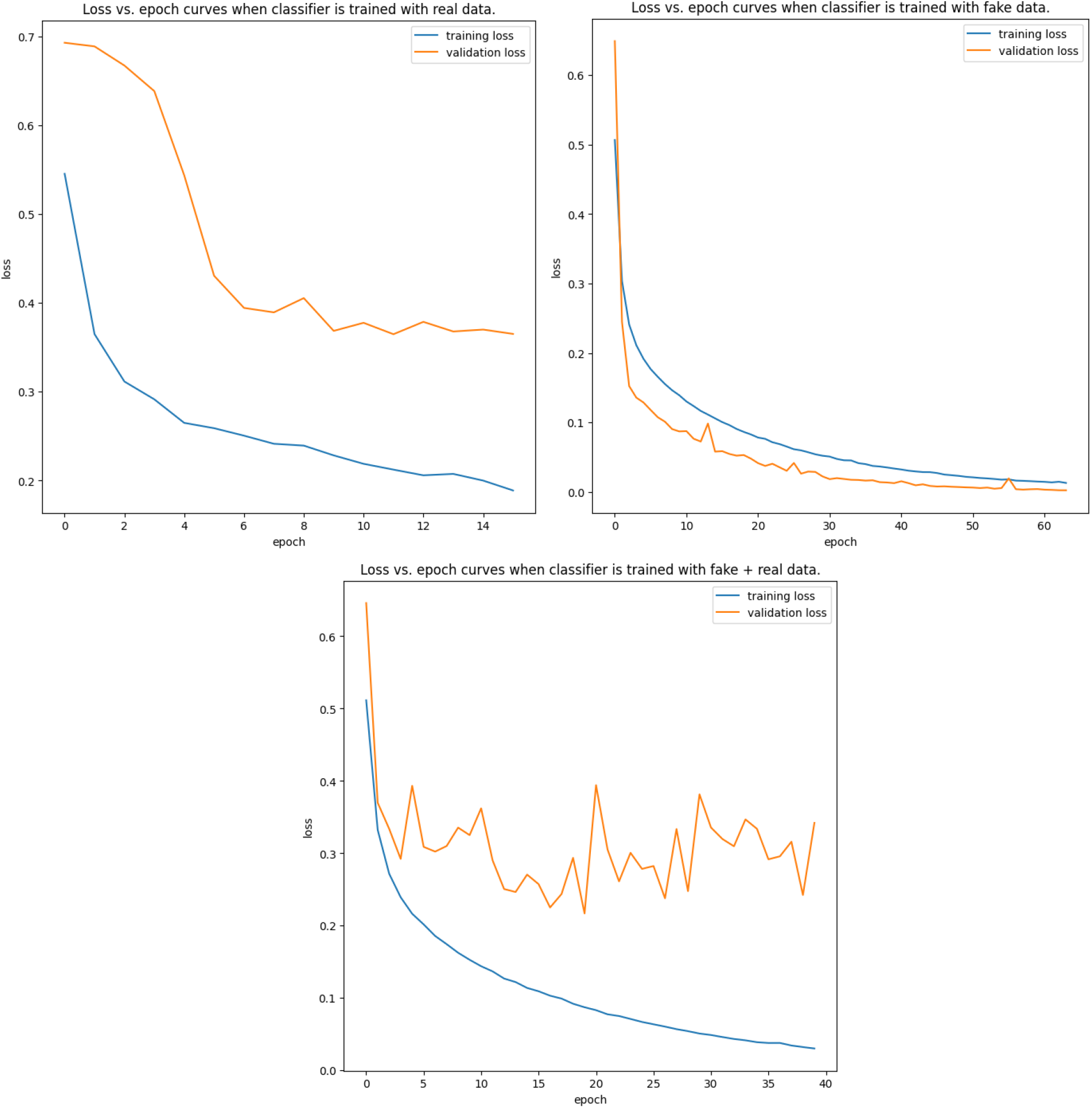
Training curves for Chest X-ray classification

## VII. Additional Experiments on brain MRI images

We perform several additional experiments along a different modality of brain MRI images by visualizing the synthetic data generated using AC-GAN, trained on the brain tumor dataset (outlined in sec. V.1). We generate images across 2 classes: ‘Positive’, and ‘Negative’, denoting tumorous and healthy MRI scans, respectively. The quantitative analysis is similar to sec. VI., where we train image classification model EfficientNet v2 and compute the metrics with the model trained on real data only, fake data only, and a mix of both fake and real data. We use the same metrics: classification accuracy, precision, recall, and F1-score, to effectively evaluate the case of class imbalance.

Figure 6 reports the visual results of images generated using our proposed AC-GAN trained on brain MRI images, belonging to the ‘Negative’ class, i.e., a healthy brain without a tumor. The generated images showcase realistic representations of brain MRI scans and exhibit characteristic features of a healthy brain. This demonstrates the ability of AC-GAN to be extended to multiple modalities and input types, at the same time, reproducing complex anatomical structures without much distortion. Similarly, for the class of ‘Positive’ (containing tumors, figure 6), the generated images display faithful features associated with the presence of a tumor across the brain’s cross-section, that too across multiple planes. There exists variation in tumor size, shape, and location, proving the effectiveness of GAN in learning diverse pathological characteristics associated with a disease.

**Figure 6.**
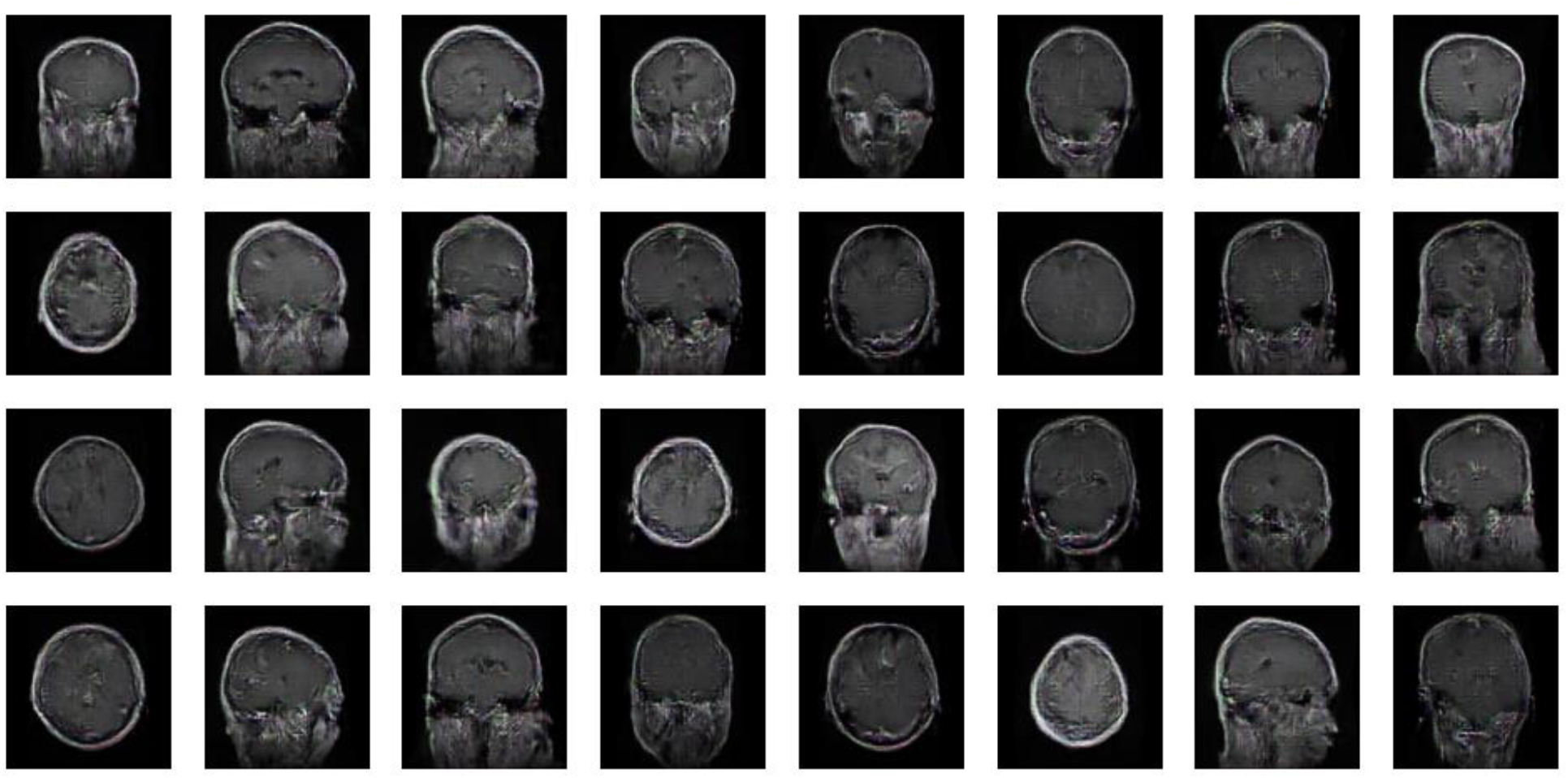
Images of class ‘Negative’ generated by proposed AC-GAN

**Figure 6.**
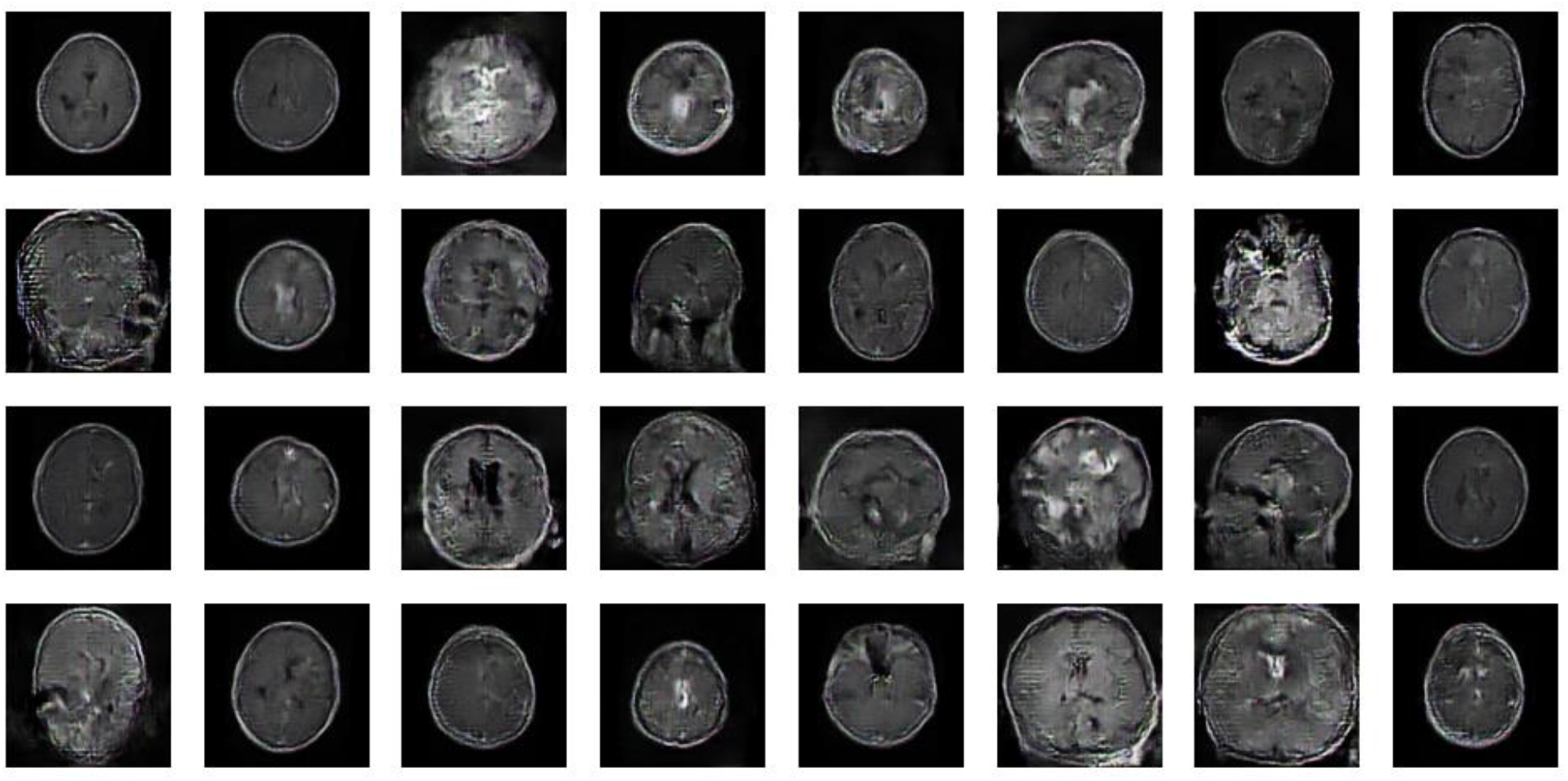
Images of class ‘Positive’ generated by proposed AC-GAN

We also perform quantitative evaluation as reported in table II. We can draw similar insights to the experiments performed in sec. V. When trained solely on real data, the model exhibits the least accuracy, precision, recall and F1-score, compared to the other two training sets. When the fake data is combined with real data to train the classifier, the model achieved highest accuracy of 85.92%, with a peak recall of 0.82 and impressive F1-score of 0.84.

**Table II.**
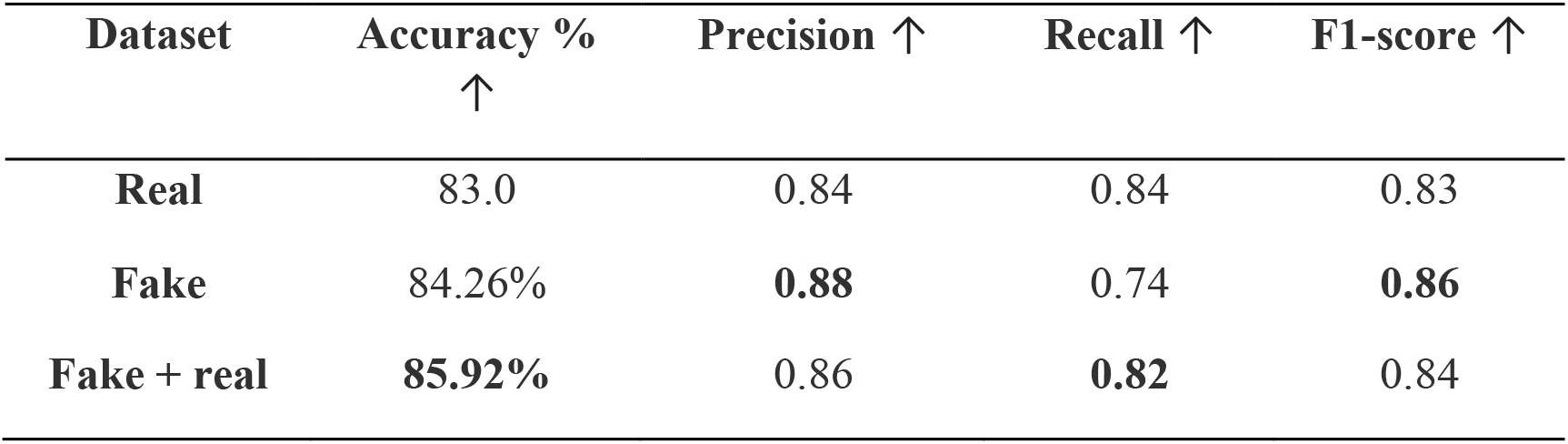
Classifier performance when trained with real, fake, and fake + real data on brain MRI classification task.

| Dataset | Accuracy %<br>↑ | Precision ↑ | Recall ↑ | F1-score ↑ |
| --- | --- | --- | --- | --- |
| **Real** | 83.0 | 0.84 | 0.84 | 0.83 |
| **Fake** | 84.26% | **0.88** | 0.74 | **0.86** |
| **Fake + real** | **85.92%** | 0.86 | **0.82** | 0.84 |

Thus, quantitative evaluation performed on the extended modality of brain MRI scans further strengthen our hypothesis of utilizing the power of generative networks like AC-GAN to perform synthetic data augmentation and boost the performance of medical image analysis tasks.

## VII. Conclusion

In conclusion, our work demonstrates the efficacy of using synthetic data augmentation, particularly through Generative Adversarial Networks (GANs), to enhance the performance of chest X-ray classification models. By generating synthetic data representing both healthy and pathological conditions, including bacterial and viral pneumonia, we overcome limitations associated with data scarcity and labor-intensive labeling processes. Results show significant improvements in classification accuracy, precision, recall, and F1-score metrics when incorporating synthetic data alongside real data. The Auxiliary Classifier GAN (AC-GAN) architecture proves effective in generating high-quality synthetic chest X-ray images, contributing to better model generalization and robustness. Furthermore, we extended our experiments by adding another modality of brain MRI scans by simply swapping the training dataset and achieved similar trends, showcasing the power of AC-GAN. This extension underscores the versatility and potential of AC-GAN in different medical imaging contexts. While promising, further validation on larger datasets and exploration of additional GAN architectures are needed. Additionally, ethical considerations regarding patient privacy and data security must be carefully addressed. Overall, synthetic data augmentation holds great potential for advancing medical image analysis, leading to improved healthcare outcomes.

## Data Availability

All data produced in the present study are available upon reasonable request to the authors.

https://www.kaggle.com/datasets/paultimothymooney/chest-xray-pneumonia

https://www.kaggle.com/datasets/praneet0327/brain-tumor-dataset

https://www.news-medical.net/health/Viral-vs-Bacterial-Pneumonia.aspx

## Notes

### Competing Interest Statement

The authors have declared no competing interest.

### Funding Statement

This study did not receive any funding

